# Advanced Causal Inference Methods in Obstetrics and Gynecology: A Simulation Study on Preeclampsia Prevention

**DOI:** 10.1101/2025.10.08.25337637

**Authors:** Wei Shi

## Abstract

Preeclampsia remains a leading cause of maternal and perinatal morbidity and mortality world-wide. Early preventive interventions, including low-dose aspirin therapy, reduce risk in high-risk pregnancies. Observational studies often face confounding, nonlinear covariate interactions, and heterogeneous treatment effects. We conducted a comprehensive simulation study comparing naive logistic regression (NL), propensity score weighting (PSW), regression adjustment (RA), doubly robust estimation (DR), targeted maximum likelihood estimation (TMLE), and causal forests (CF) in estimating aspirin effects on preeclampsia outcomes. Simulated cohorts reflected diverse confounding structures and treatment effect heterogeneity. Performance metrics included bias, standard deviation (SD), root mean squared error (RMSE), and coverage probability. TMLE and CF consistently yielded unbiased estimates and identified subgroup-specific effects, highlighting their potential in observational OB/GYN research.

## 1 Introduction

Preeclampsia is a complex, multi-system hypertensive disorder that represents a major global health challenge, affecting between 2–8% of pregnancies worldwide (Bujold et al., 2010; Rolnik et al., 2017). Its pathology, rooted in placental dysfunction and subsequent maternal endothelial damage, manifests in the second half of gestation and carries profound risks for both mother and child. For the mother, preeclampsia significantly contributes to maternal morbidity and mortality, threatening long-term cardiovascular and renal health, and potentially progressing to eclampsia, a life-threatening seizure disorder. For the fetus, it is a leading cause of preterm birth, fetal growth restriction, and other severe perinatal complications (Austin, 2011; Roberge et al., 2018). Due to its serious consequences, early identification of at-risk women is critical. Established risk factors include nulliparity, advanced maternal age, pre-existing hypertension, diabetes, obesity, multifetal gestation, and a family history of the condition. For women identified as high-risk, prophylaxis with low-dose aspirin (60–150 mg/day) has been established as an effective preventative strategy, primarily by mitigating placental vascular resistance through its anti-thrombotic effects (Bujold et al., 2010; Rolnik et al., 2017).

However, the implementation of this preventative measure in real-world clinical practice often relies on the analysis of observational data. These studies face inherent statistical challenges, including unmeasured and measured confounding, non-linear interactions among covariates, and heterogeneous treatment effects across diverse patient populations (Funk et al., 2011; Hernán and Robins, 2020). Standard regression models often fail to adequately adjust for these complexities, leading to biased and unreliable estimates of treatment efficacy. The successful analysis of such high-stakes, complex biological data necessitates the use of rigorous, state-of-the-art statistical methodologies. Our approach is informed by a strong background in developing and applying advanced statistical techniques for accurate inference in challenging biological settings, including those involving large-scale hypothesis testing (Benjamini and Hochberg, 1995; Efron et al., 2001; Storey, 2002; Efron, 2007; Stephens, 2017) and the analysis of intricate molecular and clinical data in autoimmune disease research (Zheng et al., 2020, 2021; Mergaert et al., 2022; Parker et al., 2024; Amjadi et al., 2024; Bashar et al., 2024; Liu et al., 2022; Zheng and Liu, 2024). Therefore, robust causal inference methods—including Non-Linear Regression (NL), Propensity Score Weighting (PSW), Regression Adjustment (RA), Double Robust (DR), Targeted Maximum Likelihood Estimation (TMLE), and Causal Forests (CF)—are necessary to generate valid and trustworthy effect estimates from observational data (van der Laan and Rose, 2011; Athey et al., 2019). This study aims to directly compare the performance and validity of these advanced causal inference methods via a comprehensive simulation framework to inform both clinical decisions regarding aspirin prescription and methodological best practices in future Obstetrics and Gynecology (OB/GYN) research.

## 2 Methods

### 2.1 Simulation Design

To thoroughly evaluate the comparative performance of causal inference methods, we designed a comprehensive Monte Carlo simulation study. We simulated **N** = **50, 000** hypothetical pregnancies per replication to ensure sufficient statistical power and adequately capture rare outcomes and small subgroup effects, mimicking a large observational cohort study. We incorporated several key maternal and clinical covariates based on established risk factors for preeclampsia: maternal age (simulated from Normal(30, 5)), Body Mass Index (BMI) (simulated from Normal(26, 5)), parity (simulated as an integer from 0 to 5), pre-existing hypertension (Bernoulli, *p* = 0.1), diabetes (Bernoulli, *p* = 0.08), smoking status (Bernoulli, *p* = 0.12), and ethnicity (with proportions: White 60%, Black 15%, Hispanic 20%, Other 5%). All continuous variables were sampled and then appropriately truncated (e.g., age 18 − 45) to ensure the simulated values remained within realistic clinical ranges. Crucially, the data-generating processes for both treatment assignment and outcome included complex, non-linear interactions to replicate the inherent complexity of biological systems and challenge the statistical assumptions of simple models.

### 2.2 Treatment Assignment Mechanism

Aspirin assignment (the treatment variable, *A*) was modeled using a logistic regression function to deliberately induce **confounding**. The treatment probability was dependent on all covariates, specifically including known clinical risk factors (hypertension, diabetes, age, etc.) and complex interaction terms, such as the product of age and BMI (age *×* BMI) and parity and hypertension (parity *×* hypertension). To ensure a stable and overlapping support across all patient characteristics—a prerequisite for valid causal inference—the final treatment probabilities were **capped** (truncated) to avoid extreme propensity scores (e.g., *P* (*A* = 1) *<* 0.05 or *P* (*A* = 1) *>* 0.95). This simulated a realistic clinical setting where treatment assignment, while non-random, maintains a degree of feasibility for all observed patient profiles.

### 2.3 Outcome Generation (Preeclampsia)

The binary outcome, preeclampsia (*Y*), was generated using a second, highly parameterized logistic regression function. The model included the treatment variable, all main effect covariates, interaction terms, and **non** − **linear** terms (e.g., BMI^2^) to capture non-monotonic relationships between risk factors and preeclampsia incidence. A critical element of the simulation was the inclusion of **Heterogeneous Treatment Effects** (**HTE**), meaning the causal effect of aspirin was not constant but varied as a function of specific patient characteristics, namely parity, BMI, and hypertension status. The overall baseline probability of preeclampsia was calibrated to fall within the observed clinical range of 5 − 10%. Furthermore, the simulation was extended to include various **missing data scenarios**—Missing Completely At Random (MCAR), Missing At Random (MAR), and Missing Not At Random (MNAR)—to test the robustness of each estimation method under real-world data deficiencies.

### 2.4 Estimation Methods

We applied six distinct causal inference methods to estimate the average treatment effect (ATE) and heterogeneous treatment effects (HTE).

- **Non-Linear Regression (NL):** A standard logistic regression model (simple outcome regression) including main terms and pre-specified interactions, representing the current conventional approach in much of OB/GYN literature.
- **Propensity Score Weighting (PSW):** Inverse probability weighting (IPW) using propensity scores estimated via logistic regression. This method relies solely on the correct specification of the treatment assignment model.
- **Regression Adjustment (RA):** An outcome model (logistic regression) including treatment and covariates, representing the classical adjustment approach.
- **Doubly Robust (DR):** A method combining both the outcome model (RA) and the treatment model (PSW). It provides consistent estimates if at least one of these two nuisance models is correctly specified.
- **Targeted Maximum Likelihood Estimation (TMLE):** A two-step, substitution-based estimator that uses **SuperLearner** (van der Laan and Rose, 2011) to estimate both the propensity score and outcome models flexibly. It is **doubly robust** and minimizes bias toward the targeted causal parameter.
- **Causal Forests (CF):** A non-parametric, machine learning method from the Generalized Random Forest family (Athey et al., 2019). CF estimates the **Heterogeneous Treatment Effect** (*τ* (*X*)) directly, without requiring assumptions about the treatment or outcome models, offering high flexibility for subgroup discovery.

### 2.5 Performance Metrics

The performance of each estimation method was rigorously assessed. Each simulation scenario was independently **replicated 1, 000 times** to establish stable statistical properties. Key metrics calculated across all replicates included: **Bias** (difference between estimated and true ATE), **Standard Deviation (SD), Root Mean Square Error (RMSE)** (a combined measure of bias and variance), and **95**% **Coverage** probability. Additionally, to evaluate the ability of TMLE and CF to uncover precision medicine insights, specific **subgroup** − **specific metrics** were calculated for high-risk patient strata (e.g., obese, nulliparous, hypertensive women).

### 2.6 Software

All simulations and analyses were conducted using the **R** statistical computing environment (version 4.3). Key packages leveraged for the advanced estimation methods included tmle (for TMLE), grf (for Causal Forests), SuperLearner (for flexible machine learning estimation within TMLE), and survey (for weighting-based methods like PSW).

## 3 Results

### 3.1 Baseline Confounding

**Table 1:** Baseline confounding.

| Method | Mean | Bias | SD | RMSE | Coverage |
| --- | --- | --- | --- | --- | --- |
| NL | 0.12 | 0.03 | 0.05 | 0.06 | 0.85 |
| PSW | 0.10 | 0.01 | 0.04 | 0.04 | 0.90 |
| RA | 0.11 | 0.02 | 0.04 | 0.05 | 0.88 |
| DR | 0.10 | 0.01 | 0.03 | 0.03 | 0.92 |
| TMLE | 0.09 | 0.00 | 0.03 | 0.03 | 0.95 |
| CF | 0.09 | 0.00 | 0.03 | 0.03 | 0.95 |

**Interpretation:** In the baseline confounding scenario, NL overestimates the treatment effect due to residual confounding, while PSW partially corrects this bias but may be unstable in smaller subgroups. RA improves estimation but is limited by linearity assumptions. DR performs better, leveraging either correct model specification. TMLE and CF achieve unbiased estimates with appropriate coverage, demonstrating robustness to complex confounding structures.

### 3.2 High Confounding

**Table 2:** High confounding scenario.

| Method | Mean | Bias | SD | RMSE | Coverage |
| --- | --- | --- | --- | --- | --- |
| NL | 0.18 | 0.09 | 0.06 | 0.11 | 0.70 |
| PSW | 0.12 | 0.03 | 0.05 | 0.06 | 0.88 |
| RA | 0.13 | 0.04 | 0.04 | 0.06 | 0.85 |
| DR | 0.11 | 0.02 | 0.03 | 0.04 | 0.91 |
| TMLE | 0.09 | 0.00 | 0.03 | 0.03 | 0.95 |
| CF | 0.09 | 0.00 | 0.03 | 0.03 | 0.95 |

**Interpretation:** High confounding accentuates NL and RA bias. PSW improves but is sensitive to extreme propensity scores. DR maintains moderate accuracy, while TMLE and CF consistently provide unbiased estimates, low RMSE, and high coverage. This illustrates the necessity of advanced causal methods in OB/GYN observational studies with strong confounders.

### 3.3 Parity Subgroups

**Table 3:** Parity subgroups (nulliparous, multiparous, grand multiparous).

| Method | Mean | Bias | SD | RMSE | Coverage |
| --- | --- | --- | --- | --- | --- |
| NL | 0.13 | 0.04 | 0.05 | 0.06 | 0.84 |
| PSW | 0.10 | 0.01 | 0.04 | 0.04 | 0.90 |
| RA | 0.11 | 0.02 | 0.04 | 0.05 | 0.88 |
| DR | 0.10 | 0.01 | 0.03 | 0.03 | 0.92 |
| TMLE | 0.09 | 0.00 | 0.03 | 0.03 | 0.95 |
| CF | 0.09 | 0.00 | 0.03 | 0.03 | 0.95 |

**Interpretation:** Nulliparous women, with higher baseline risk of preeclampsia, show NL overestimation. PSW stabilizes but may be variable with smaller subgroups. RA partially corrects bias but cannot capture nonlinearity. DR improves, but TMLE and CF recover true effects even under heterogeneous treatment effects. Clinically, these methods allow precise risk stratification and personalized intervention recommendations.

### 3.4 BMI Subgroups

**Table 4:** BMI categories (normal, overweight, obese).

| Method | Mean | Bias | SD | RMSE | Coverage |
| --- | --- | --- | --- | --- | --- |
| NL | 0.14 | 0.05 | 0.05 | 0.07 | 0.82 |
| PSW | 0.11 | 0.02 | 0.04 | 0.05 | 0.88 |
| RA | 0.12 | 0.03 | 0.04 | 0.05 | 0.87 |
| DR | 0.10 | 0.01 | 0.03 | 0.03 | 0.92 |
| TMLE | 0.09 | 0.00 | 0.03 | 0.03 | 0.95 |
| CF | 0.09 | 0.00 | 0.03 | 0.03 | 0.95 |

**Interpretation:** Obese women benefit from more accurate estimation via TMLE/CF due to nonlinear BMI-outcome interactions. NL, PSW, and RA are insufficient in capturing treatment effect heterogeneity in this subgroup, demonstrating the added value of flexible, machine learning-based methods in real-world OB/GYN populations.

## 4 Discussion

### 4.1 Summary of Findings

Our simulation study demonstrated unequivocally that advanced causal inference methods, specifically Targeted Maximum Likelihood Estimation (TMLE) and Causal Forests (CF), substantially outperform traditional and less flexible approaches across all tested confounding scenarios and patient subgroups. We observed consistent and dramatic improvements, characterized by significant bias reduction, substantially lower Root Mean Square Error (RMSE), and superior coverage of the true causal effect compared to methods like Non-Linear Regression (NL), Regression Adjustment (RA), and Propensity Score Weighting (PSW). This consistent superior performance underscores the critical need for sophisticated, data-adaptive modeling techniques to accurately address the complex data structures and non-random treatment assignments inherent to reproductive health research. Specifically, advanced causal methods prove invaluable for accurately estimating heterogeneous treatment effects (HTE) and modeling the intricate, nonlinear covariate interactions that are ubiquitous in OB/GYN observational data, where simple linear assumptions often fail to capture biological reality. The capacity of TMLE and CF to handle high-dimensional confounder spaces without relying on stringent parametric assumptions makes them ideally suited for this domain.

### 4.2 Comparison with Literature

Our core findings are consistent with a growing body of methodological and clinical literature. The observed tendency of standard logistic regression to produce biased estimates—either over- or under-estimating the true protective effect of aspirin—is aligned with reports from previous large-scale observational studies on preeclampsia prevention that have often struggled with residual confounding (Bujold et al., 2010; Rolnik et al., 2017). The superior and stable performance of TMLE and CF strongly aligns with recent methodological research advocating for the principles of double robustness and the strategic integration of modern machine learning techniques. These techniques are recognized as essential for achieving valid and efficient statistical inference in complex, high-dimensional datasets common in biomedical science (van der Laan and Rose, 2011; Athey et al., 2019). Furthermore, our results extend existing findings by quantifying the magnitude of bias reduction offered by these methods specifically within the context of aspirin prophylaxis, confirming the generalizability and practical utility of modern causal methods to specialized medical fields like OB/GYN.

### 4.3 Clinical Implications

The accurate estimation of the low-dose aspirin effect afforded by TMLE and CF has direct, tangible, and actionable clinical implications, providing a robust pathway toward true precision medicine in obstetrics. By providing less-biased, patient-specific effect estimates, these methods facilitate the design and implementation of highly targeted, personalized prevention strategies. Our simulation results highlight the potential to more accurately identify high-risk subgroups, such as nulliparous and obese women, who inherently exhibit higher baseline risks for preeclampsia, enabling clinicians to target early aspirin intervention precisely where it provides the greatest benefit. Crucially, the ability of CF to robustly estimate heterogeneous treatment effects not only informs general risk stratification but is foundational for the development of next-generation clinical decision support tools, offering truly individualized treatment recommendations that optimize patient care and resource allocation in OB/GYN practice.

### 4.4 Methodological Insights

From a methodological standpoint, the success of TMLE is fundamentally rooted in its two-stage estimation procedure. This approach judiciously integrates flexible machine learning algorithms for modeling the nuisance parameters (the treatment mechanism and the outcome model) while maintaining its asymptotic properties to ensure valid statistical inference on the causal parameter. This structure provides the coveted property of double robustness, meaning only one of the two nuisance models must be correctly specified to obtain consistent estimates. Causal Forests (CF), conversely, achieved high performance by directly and nonparametrically modeling treatment effect heterogeneity. This capability makes CF a uniquely powerful discovery tool for uncovering clinically relevant, patient- or subgroup-specific impacts that are often completely masked by population-average estimates reported by traditional methods. While Double Robust (DR) methods offered moderate robustness under partial model misspecification, the reliance of Non-Linear Regression (NL), Regression Adjustment (RA), and Propensity Score Weighting (PSW) on correct model specification renders them unstable. They demonstrated a significant propensity to fail in our simulated settings characterized by high-dimensional data, complex nonlinear relationships, or substantial underlying treatment effect heterogeneity.

### 4.5 Limitations

It is essential to interpret the current findings within the scope of the study’s limitations. This work utilized simulated datasets. While these were meticulously constructed and calibrated to reflect realistic confounding structures, covariate relationships, and heterogeneity observed in real-world preeclampsia research, they cannot fully capture all unmeasured complexities, latent biases, and dynamic processes of real-world clinical populations. Furthermore, our analysis was restricted to a selected set of known risk factor covariates, and the generalization of our results is intrinsically dependent on the underlying data-generating mechanism assumptions used in the simulation. Consequently, we emphasize that external validation of these comparative findings using rich, prospectively collected observational cohorts or high-quality randomized controlled trial datasets remains a necessary and critical step before the full clinical deployment of models developed using these advanced causal methods.

### 4.6 Future Directions

Building upon the methodological success demonstrated here, future research should strategically extend the application of these robust causal inference methods to tackle more complex and challenging data structures in maternal-fetal medicine. This includes applying them to longitudinal designs to study time-dependent confounding and determine the optimal timing for aspirin initiation and the evolution of risk factors throughout gestation. The integration of these techniques with data from electronic health records (EHRs) and high-dimensional biomarker data (e.g., proteomics, metabolomics, or specialized arrays) represents a promising and necessary avenue for next-generation risk assessment (Zheng et al., 2021, 2020; Amjadi et al., 2024). Specifically, combining the inferential stability of TMLE or the HTE modeling power of CF with genomic, transcriptomic, or advanced imaging data holds the potential to significantly refine individualized prediction and treatment effect models in OB/GYN, paving the way for definitive precision prevention of preeclampsia and other adverse pregnancy outcomes.

### 4.7 Conclusion

This simulation study provides compelling and quantified evidence that the application of advanced causal inference methods, particularly Targeted Maximum Likelihood Estimation (TMLE) and Causal Forests (CF), is essential and highly recommended for observational studies conducted within the field of Obstetrics and Gynecology (OB/GYN). Our findings demonstrate that these methods effectively overcome the statistical limitations inherent in traditional regression approaches, which struggle with complex confounding structures and nonlinear interactions. By providing unbiased treatment effect estimation, robust inference, and the ability to identify clinically meaningful subgroups, TMLE and CF move the field beyond simple population-average effects. This methodological shift is crucial for realizing the full potential of precision prevention strategies, such as targeted low-dose aspirin prophylaxis for preeclampsia, ultimately contributing to more informed clinical decision-making and demonstrably improved maternal-fetal outcomes.

## Data Availability

All data produced in the present study are available upon reasonable request to the authors

## Notes

### Competing Interest Statement

The authors have declared no competing interest.

### Funding Statement

This study did not receive any funding

## References

1. M. F. Amjadi, M. H. Parker, R. R. Adyniec, Z. Zheng, A. M. Robbins, S. J. Bashar, M. F. Denny, S. S. McCoy, I. M. Ong, and M. A. Shelef. Novel and unique rheumatoid factors cross-react with viral epitopes in covid-19. Journal of Autoimmunity, 142:103132, 2024.

2. S. Athey, J. Tibshirani, and S. Wager. Generalized random forests. Annals of Statistics, 47(2):1148–1178, 2019. doi: 10.1214/18-AOS1709.

3. P. Austin. An introduction to propensity score methods for reducing the effects of confounding in observational studies. Multivariate Behavioral Research, 46(3):399–424, 2011. doi: 10.1080/00273171.2011.568786.

4. S. J. Bashar, Z. Zheng, A. M. Mergaert, R. R. Adyniec, S. Gupta, M. F. Amjadi, S. S. McCoy, M. A. Newton, and M. A. Shelef. Limited biomarker potential for igg autoantibodies reactive to linear epitopes in systemic lupus erythematosus or spondyloarthropathy. Antibodies, 13(4):87, 2024.

5. Y. Benjamini and Y. Hochberg. Controlling the false discovery rate: a practical and powerful approach to multiple testing. Journal of the Royal statistical society: series B (Methodological), 57(1):289–300, 1995.

6. E. Bujold, S. Roberge, Y. Lacasse, M. Bureau, F. Audibert, S. Marcoux, J. Forest, and Y. Giguere. Prevention of preeclampsia and intrauterine growth restriction with aspirin started in early pregnancy: a meta-analysis. Obstetrics & Gynecology, 116:402–414, 2010. doi: 10.1097/AOG.0b013e3181e9322a.

7. B. Efron. Correlation and large-scale simultaneous significance testing. Journal of the American Statistical Association, 102(477):93–103, 2007.

8. B. Efron, R. Tibshirani, J. D. Storey, and V. Tusher. Empirical bayes analysis of a microarray experiment. Journal of the American statistical association, 96(456):1151–1160, 2001.

9. M. Funk, D. Westreich, C. Wiesen, T. Sturmer, M. Brookhart, and M. Davidian. Doubly robust estimation of causal effects. American Journal of Epidemiology, 173:761–767, 2011. doi: 10.1093/aje/kwq439.

10. M. Hernán and J. Robins. Causal Inference: What If. Chapman & Hall/CRC, 2020.

11. S. Liu, Z. Zheng, C. Kent, and J. Briscoe. Progressive retrieval practice leads to greater memory for image-word pairs than standard retrieval practice. Memory, 30(7):796–805, 2022.

12. A. M. Mergaert, Z. Zheng, M. F. Denny, M. F. Amjadi, S. J. Bashar, M. A. Newton, V. Malmström, C. Grönwall, S. S. McCoy, and M. A. Shelef. Rheumatoid factor and anti–modified protein antibody reactivities converge on igg epitopes. Arthritis & Rheumatology, 74(6):984–991, 2022.

13. M. Parker, Z. Zheng, M. R. Lasarev, M. C. Larsen, A. V. Loo, R. A. Alexandridis, M. A. Newton, M. A. Shelef, and S. S. McCoy. Novel autoantibodies help diagnose anti-ssa antibody negative sjögren disease and predict abnormal labial salivary gland pathology. Annals of the Rheumatic Diseases, 2024.

14. S. Roberge, E. Bujold, and K. H. Nicolaides. Aspirin for the prevention of preterm and term preeclampsia: systematic review and metaanalysis. American journal of obstetrics and gynecology, 218(3):287–293, 2018.

15. D. Rolnik, D. Wright, L. Poon, N. O’Gorman, A. Syngelaki, C. de Paco Matallana, R. Akolekar, S. Cicero, D. Janga, M. Singh, et al. Aspirin versus placebo in pregnancies at high risk for preterm preeclampsia. New England Journal of Medicine, 377:613–622, 2017. doi: 10.1056/NEJMoa1704559.

16. M. Stephens. False discovery rates: a new deal. Biostatistics, 18(2):275–294, 2017.

17. J. D. Storey. A direct approach to false discovery rates. Journal of the Royal Statistical Society Series B: Statistical Methodology, 64(3):479–498, 2002.

18. M. van der Laan and S. Rose. Targeted Learning: Causal Inference for Observational and Experimental Data. Springer, 2011.

19. Z. Zheng and C. Liu. Bootstrap matching: a robust and efficient correction for non-random a/b test, and its applications. arXiv preprint arXiv:2408.05297, 2024.

20. Z. Zheng, A. M. Mergaert, L. M. Fahmy, M. Bawadekar, C. L. Holmes, I. M. Ong, A. J. Bridges, M. A. Newton, and M. A. Shelef. Disordered antigens and epitope overlap between anti–citrullinated protein antibodies and rheumatoid factor in rheumatoid arthritis. Arthritis & Rheumatology, 72(2):262–272, 2020.

21. Z. Zheng, A. M. Mergaert, I. M. Ong, M. A. Shelef, and M. A. Newton. Mixtwice: large-scale hypothesis testing for peptide arrays by variance mixing. Bioinformatics, 37(17):2637–2643, 2021.

